# Dynamic functional connectivity analysis reveals transiently increased segregation in patients with severe stroke

**DOI:** 10.1101/2020.06.01.20119263

**Authors:** Anna K. Bonkhoff, Markus D. Schirmer, Martin Bretzner, Mark Etherton, Kathleen Donahue, Carissa Tuozzo, Marco Nardin, Anne-Katrin Giese, Ona Wu, Vince Calhoun, Christian Grefkes, Natalia S. Rost

## Abstract

**Background and Purpose:** To explore the whole-brain dynamic functional network connectivity patterns in acute ischemic stroke (AIS) patients and their relation to stroke severity in the short and long term.

**Methods:** We investigated large-scale dynamic functional network connectivity of 41 AIS patients two to five days after symptom onset. Re-occurring dynamic connectivity configurations were obtained using a sliding window approach and k-means clustering. We evaluated differences in dynamic patterns between three NIHSS-stroke severity defined groups (mildly, moderately, and severely affected patients). Furthermore, we established correlation analyses between dynamic connectivity estimates and AIS severity as well as neurological recovery within the first 90 days after stroke (DNIHSS). Finally, we built Bayesian hierarchical models to predict acute ischemic stroke severity and examine the inter-relation of dynamic connectivity and clinical measures, with an emphasis on white matter hyperintensity lesion load.

**Results:** We identified three distinct dynamic connectivity configurations in the early post-acute stroke phase. More severely affected patients (NIHSS 10–21) spent significantly more time in a highly segregated dynamic connectivity configuration that was characterized by particularly strong connectivity (three-level ANOVA: *p*<0.05, post hoc t-tests: *p*<0.05, FDR-corrected for multiple comparisons). Recovery, as indexed by the realized change of the NIHSS over time, was significantly linked to the acute dynamic connectivity between bilateral intraparietal lobule and left angular gyrus (Pearson’s *r* = –0.68, *p*<0.05, FDR-corrected). Increasing dwell times, particularly those in a very segregated connectivity configuration, predicted higher acute stroke severity in our Bayesian modelling framework.

**Conclusions:** Our findings demonstrate transiently increased segregation between multiple functional domains in case of severe AIS. Dynamic connectivity involving default mode network components significantly correlated with recovery in the first three months post-stroke.

## Introduction

One in four adults over the age of 25 experiences a stroke during their lifetime^1^ and is thus frequently confronted with long term impairments in multiple functional domains^2^. Establishing a comprehensive understanding of cerebral changes early after stroke is of prime importance to successfully design rehabilitative strategies. In this respect, functional neuroimaging has proven to be an instrumental tool to uncover neural mechanisms of plasticity and reorganization post-stroke^3,4^. Motor impairments have previously been linked to distinct decreases in functional connectivity between sensorimotor areas within and across hemispheres^5,6,7,8^. Conversely, post-stroke cognitive impairments and affective symptoms, such as anxiety and depression, have been associated with functional connectivity alterations in default mode network regions^9,10^.

Conventional resting-state functional MRI (rsfMRI) analyses typically evaluate functional connectivity over the duration of entire scan sessions (i.e., several minutes). Contrariwise, the recently developed time-varying – *dynamic* – functional network connectivity (dFNC) analyses allow connectivity strengths to fluctuate from moment to moment and, by these means, enable a time-resolution in the order of seconds^11,12,13^. Numerous studies suggest that this dynamic approach represents a powerful tool to gain novel insights into brain connectivity changes associated with to neurological diseases, e.g., migraine^14^, Parkinson’s disease^15,16^, and Huntington’s disease^17^. In case of ischemic stroke, the dFNC analysis of motor domains has been essential in uncovering transiently increased integration in stroke patients with moderate motor impairment and a significant preference for segregated dynamic connectivity states in patients with severe motor impairments^18^.

In extension to previous dFNC analyses in acute ischemic stroke (AIS), we here investigated dynamic connectivity not only in motor domains, but within the entire brain of 41 AIS patients. We evaluated links between dynamic connectivity estimates and acute, as well as chronic stroke severity three months post-stroke and hypothesized that large-scale, transient alterations in functional integration and segregation were driven by differences in AIS severity. Furthermore, we examined whether dynamic connectivity derived estimates augmented the prediction of AIS severity beyond the prediction relying on clinical information, such as the white matter hyperintensity (WMH) lesion load, only.

## Materials and Methods

### Participants

We considered rsfMRI scans from forty-seven AIS patients who were admitted to the Emergency Department at Massachusetts General Hospital, USA and subsequently enrolled in the SALVO (statins augment small vessel function and improve stroke outcomes) study between 2014 and 2019. The following inclusion criteria were applied: (i) admission ≤24 hours from onset of focal neurological symptoms consistent with a cerebrovascular syndrome, (ii) MRI findings corresponding to acute cerebral ischemic injury (e.g., DWI-positive lesions), and (iii) evidence of moderate to severe WMH lesion load (Fazekas grade ≥2^19^). Exclusion criteria included: (i) the evidence of secondary cause of stroke or primary hemorrhagic stroke, (ii) medical contraindications to MRI or gadolinium-based contrast agents, (iii) the inability to complete the study due to severe medical or behavioral co-morbidities, as determined by PI during screening, and (iv) pregNancy or lactation at the time of screening. One stroke patient had to be excluded due to structural abnormalities (benign cyst located in the temporal lobe) and five further participants due to pronounced head motion during image acquisition (c.f., below). Therefore, 41 subjects were included in the analyses. The study was approved by the institutional review board (Massachusetts General Hospital) and all participants, or their surrogates, gave written informed consent at the time of enrollment.

### Clinical Assessment

Individual stroke severity, quantified by means of the National Institutes of Health Stroke Scale score (NIHSS), and functional outcome, assessed by the modified Rankin Scale score (mRS), were obtained by trained neurologists at multiple instances post-stroke (admission, time of the rsfMRI scan, 90day). We defined three groups of stroke patients based on NIHSS scores at time of scanning: mildly affected (NIHSS: 0–2, n = 19), moderately affected (NIHSS: 3–9, n = 15), and severely affected (NIHSS 10–21, n = 7). Cut-offs were chosen based on the sample-specific distribution of NIHSS scores with the aim of creating as homogeneous groups with regard to stroke severity as possible. In view of the less frequent recruitment of severely affected patients and resulting group imbalance, we additionally performed correlation and regression analyses to supplement group-dependent with group-independent results.

## Data Acquisition

### Magnetic resonance imaging data acquisition

Patients were scanned at admission (DWI, 1.5T, General Electric scanner) as well as two to five days after stroke onset (functional scans, 3.0T, Siemens Skyra scanner). We here relied on diffusion weighted images (DWI) and rsfMRI data with the following parameters: **DWI:** echo-planar imaging, number of slices: 28; slice thickness: 5 mm; repetition time (TR): 5500 ms; echo time (TE): 99ms; in-plane resolution: 1.375 mm, and **rsfMRI:** gradient-echo planar imaging (EPI) sequence, 150 volumes, number of slices: 42 (interleaved); slice thickness: 3.51 mm; matrix size: 64×64; flip angle: 90°; repetition time (TR): 2400 ms; in-plane resolution: 3.437 mm. The entire rsfMRI scan time amounted to 6 minutes.

### Preprocessing of functional resting-state images

Preprocessing of the rsfMRI data was conducted using MATLAB (Version R2019b, MathWorks, Inc., Natick, MA, USA) and the Statistical Parametric Mapping software package (SPM12, Wellcome Trust Centre for Neuroimaging, London, UK; https://www.fil.ion.ucl.ac.uk/spm). After removal of the first three scans (“dummy images”) to ensure a steady blood oxygenation level dependent (BOLD) activity signal, the remaining 147 volumes were head movement-corrected through affine realignment to each scan’s mean image. Diffusion-weighted images, as well as corresponding binary lesion masks were linearly co-registered to the same functional mean image (c.f. lesion overlap in **Supplementary Figure 1**). All volumes were then non-linearly spatially normalized employing the “unified segmentation” option with masked lesions^20^. Finally, images were smoothed using a Gaussian kernel with a full-width at half maximum (FWHM) of 8 mm.

### Intrinsic connectivity networks

We employed the spatially constrained independent component analysis (ICA) algorithm^21,22^ to back-reconstruct individual spatial information and time-courses of 50 ICA components (as available from ref. ^12^). One component (caudate) did not pass quality control after back-reconstruction (spatial inaccuracies) and was therefore excluded. The remaining 49 components were organized into seven functional domains: i) sensorimotor, ii) subcortical, iii) cerebellar, iv) visual, v) auditory, vi) cognitive control, and vii) default mode network (**Figure 1**). Additional post-processing steps were applied: We detrended (i.e., accounted for linear, quadratic and cubic trends in the data), despiked using 3Ddespike^23^, and filtered time courses by a fifth-order Butterworth low-pass filter with a high-frequency cutoff of 0.15 Hz. Lastly, each time-course was variance-normalized^24^.

**Figure 1.**
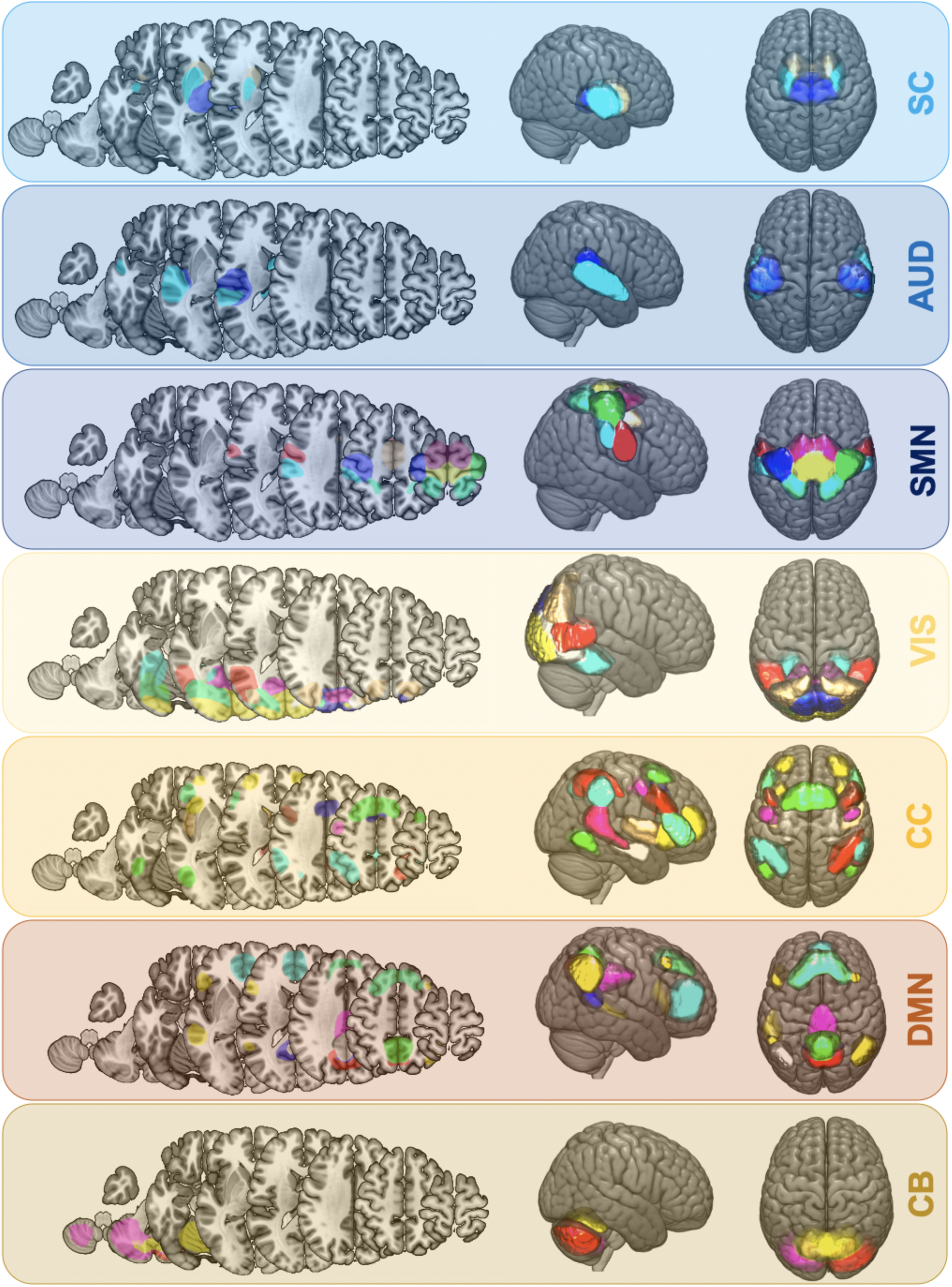
Spatial maps of 49 intrinsic connectivity networks of all ischemic stroke subjects (n = 41). Networks were assigned to seven functional domains: Subcortical (SC, 3 networks, *light blue*), auditory (AUD, 3 networks, *blue*), cortical sensorimotor (SMN, 8 networks, *dark blue*), visual (VIS, 10 networks, *yellow*), cognitive control (CC, 14 networks, *orange*), default mode network (DMN, 9 networks, *brown*), cerebellar domain (CB, 3 networks, *moccasin*). Networks were obtained via back-reconstruction of independent analysis components as given in in Allen *et al*., 2014.

### Dynamic functional network connectivity

Dynamic functional network connectivity was computed within the sliding window framework as implemented in the GIFT toolbox^12,25,26,13^. As in previous studies^12,18^, the width of the window was chosen to be close to 44 seconds (width = 18 TRs, 43.2 s). Time windows were additionally convolved with a Gaussian of seven seconds (σ = 3 TRs) and shifted at a rate of 1 TR at a time. Therefore, we obtained 129 individual tapered time windows per subjects. Dynamic connectivity between the various networks for each time window was computed via the *l*_1_-regularized precision matrix (i.e., sparse inverse covariance matrix^27,28,29^). We regressed out the covariates age, sex, mean framewise translation and rotation to decrease differences between patients. Finally, functional connectivity values were Fisher Z-transformed.

### Estimation of connectivity states

We utilized k-means clustering (*l_1_* distance^30,31^) to reveal latent dynamic functional connectivity configurations, i.e., *connectivity states*. These states represented repeating connectivity patterns across time and subjects^32,12,13^. The clustering procedure was conducted in a two-step procedure: We first derived the optimal number of clusters *k* (= number of connectivity states) in an initial run. The optimal number *k* was defined based on commonly used criteria in previous dynamic connectivity studies: The elbow criterion resting upon the cluster validity index (ratio between within-cluster distance to between-cluster distances^12^) and a state frequency of > 10%^33^. Secondly, we computed the final *k* connectivity states. For every subject, each time window was assigned to one of the *k* connectivity states. Importantly, subjects were not constraint to enter every single one of the connectivity states, which could result in fewer than the maximum number of states per subject. In a final step, we computed the following dynamic connectivity specific estimates: i) *fraction times* (the subject-specific fraction of total time spent in a state), and ii) *dwell times* (the subject-specific average time spent in a state without interruption at any one time). Additionally, we obtained subject- and state-specific dynamic connectivity strength matrices.

### Statistical analysis

#### Group differences

Differences in dynamic connectivity patterns (fraction and dwell times, dynamic connectivity strengths) with respect to group membership and thus stroke severity were determined in three-level one-way ANOVAs (mildly vs. moderately vs. severely affected patients, level of significance *p*<0.05). In case of significant ANOVA effects, we followed up with post hoc t-tests between the individual groups (mild vs. moderate, mild vs. severe, moderate vs. severe; level of significance, *p*<0.05, False discovery rate (FDR)-corrected). We then conducted correlation analyses between the significant dynamic connectivity estimates and NIHSS scores at the time of scanning (NIHSS_scan,_ Pearson correlation, level of significance, *p*<0.05, FDR-corrected). We furthermore ran partial correlation analyses between the significant dynamic connectivity estimates and NIHSS-based recovery. In analogy to a previous study^34^, we captured recovery as realized recovery potential: (NIHSS_90days_ – NIHSS_scan_)/(0 – NIHSS_scan_) (Pearson correlation, level of significance, *p*<0.05, FDR-corrected, adjusted for NIHSS_scan_). Given the inclusion criterion of a higher WMH lesion load, we performed ancillary correlation analyses between dynamic connectivity estimates and the WMH lesion load (Pearson correlation, level of significance, *p*<0.05, FDR-corrected).

#### Bayesian prediction of AIS severity

We constructed linear regression models to test the capacity of clinical and dynamic connectivity derived variables to predict stroke severity at time of the scan. We employed Bayesian hierarchical models, primarily as these models allow for i) a full estimation of parameter uncertainty (rather than point estimates), ii) an intuitive interpretation of credibility intervals and iii) Bayesian model comparisons^35^. Furthermore, given the inclusion criterion of a pronounced WMH load, the hierarchical nature of models permitted to investigate differential effects of moderate versus high loads of WMH lesions on stroke severity. The two groups of WMH load were integrated as a varying intercept. Therefore, we initiated one intercept for the moderate and one for the high lesion load group. Both of these intercepts were combined by a joint hyperprior. We then built individual models relying on either admission NIHSS (the acute baseline model, c.f. ref.^36^ for a comparable approach) or dynamic estimates (dwell times for State 1–3, the acute dynamic model). We did not integrate fraction times in addition to dwell times to reduce probable collinearity of input variables. Subsequently, we combined all of these input variables for an extended model of acute stroke severity (the acute extended model). A final Bayesian model comparison based on leave-one-out-cross-validation (LOOCV) was performed to determine the most suitable among the three models to simulate predictions of stroke severity of future patients. By means of the LOOCV, we could also account for varying numbers of input variables that can lead to too flexible models, overfitting and inflated values of explained variance. In ancillary analyses, we evaluated the effects of further covariates, i.e. age, sex and the administration of intravenous thrombolysis.

### Data and code availability

Preprocessing of data and dFNC computation used adjusted template scripts which are part of the GIFT toolbox (https://trendscenter.org/software/gift/). Additional computations were run in Python (3.7), particularly relying on the package pymc3 for Bayesian analyses^37^. Scripts will be made available upon publication [github-link].

## Results

### Clinical characteristics

Forty-one included AIS patients were scanned upon admission and two to five days after stroke onset. Stroke severity was captured at several time instances, mean NIHSS at admission was 7.8 (standard deviation (SD): 5.8, 40 patients), mean NIHSS at time of scanning: 4.7 (SD: 5.2, 41 patients) and at the 3-months follow-up: 1.3 (SD: 2.1, 29 patients). Further demographical and clinical characteristics are displayed in **Table 1**.

**Table 1.**
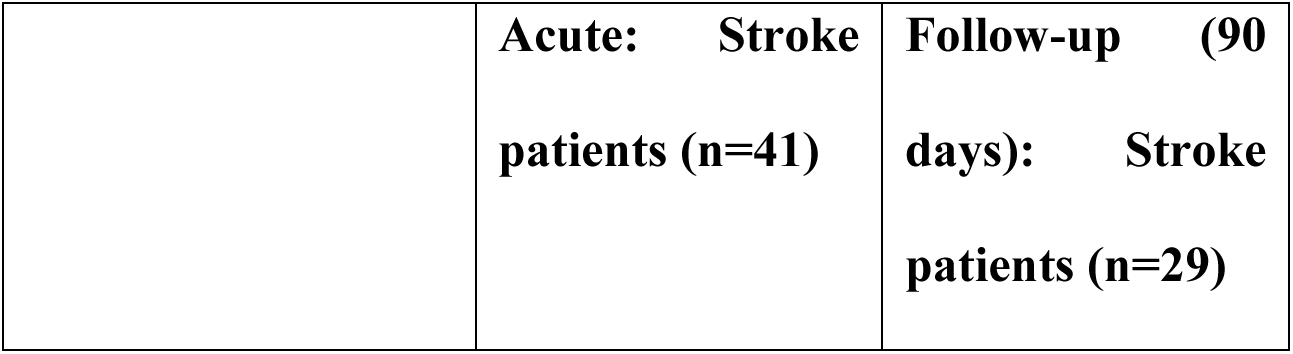
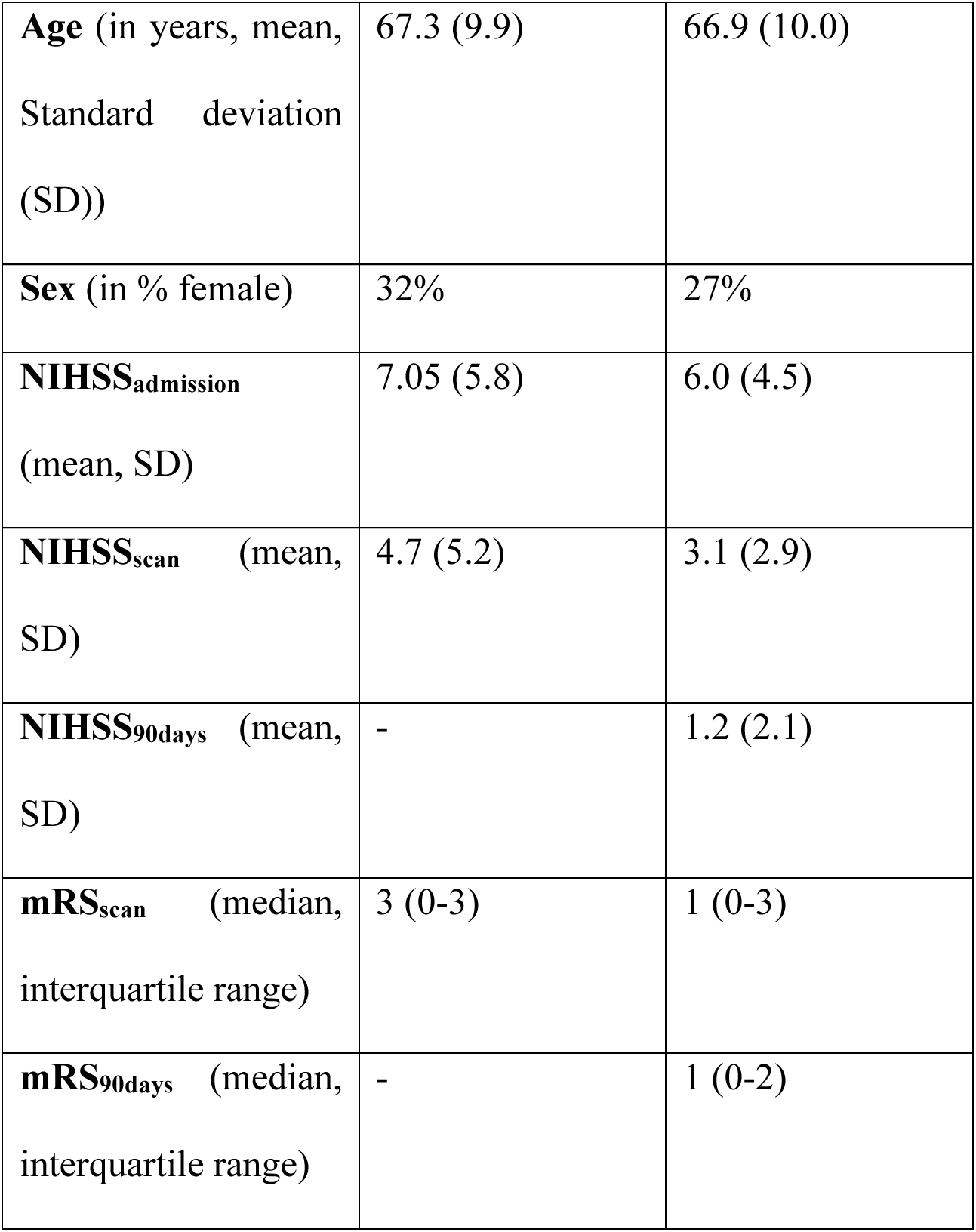
Demographics and clinical characteristics of AIS patients undergoing rsfMRI.

|  | **Acute: Stroke patients (n=41)** | **Follow-up (90 days): Stroke patients (n=29)** |
| --- | --- | --- |
| **Age** (in years, mean, Standard deviation (SD)) | 67.3 (9.9) | 66.9 (10.0) |
| **Sex** (in % female) | 32% | 27% |
| **NIHSS<sub>admission</sub>** (mean, SD) | 7.05 (5.8) | 6.0 (4.5) |
| **NIHSS<sub>scan</sub>** (mean, SD) | 4.7 (5.2) | 3.1 (2.9) |
| **NIHSS<sub>90days</sub>** (mean, SD) | - | 1.2 (2.1) |
| **mRS<sub>scan</sub>** (median, interquartile range) | 3 (0-3) | 1 (0-3) |
| **mRS<sub>90days</sub>** (median, interquartile range) | - | 1 (0-2) |

### Dynamic functional network connectivity

After the back-reconstruction of spatial maps and time courses of 49 networks originating from seven functional domains, we computed and explored characteristics of time-varying or *dynamic* functional connectivity, i.e., dFNC. Importantly, we regressed out effects of age, sex and motion at this point. The cluster validity index indicated three clusters as the optimal solution. Thus, each subject’s 129 functional connectivity matrices were assigned to one of three connectivity states (**Figure 2**).

State 1 was the most segregated state and also appeared the least often (frequency across all subjects: 21%, **Figure 2**, *left panel*). It featured highly positive intra-domain connectivity. This was particularly true for the sensorimotor, the cerebellar and visual domains. Highly negative inter-domain connectivity emerged between the cortical sensorimotor and the cerebellar as well as the visual domains. Furthermore, inter-domain connectivity between the visual and cognitive control domains was mostly highly negative. State 2, the most frequent connectivity state, displayed a markedly different connectivity pattern: It was characterized by mostly neutral inter-domain and only slightly positive intra-domain connectivity (frequency: 54%, **Figure 2**, *middle panel*). State 3, occuring with a frequency of 25%, displayed highly positive intra-domain connectivity in the sensorimotor and visual domains, resembling State 1 in this respect, but also positive inter-domain connectivity between these two domains (**Figure 2**, *right panel*).

**Figure 2.**
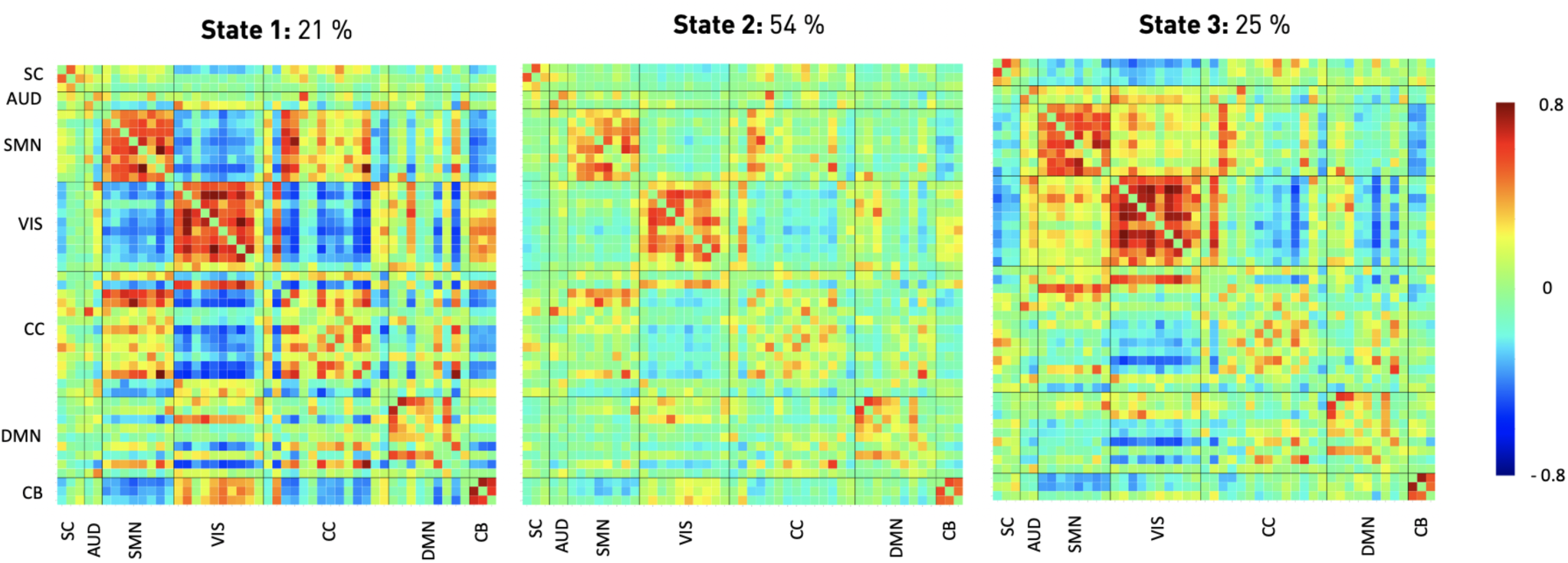
Three discrete connectivity states representing re-occurring dynamic connectivity across time and subject space. These states demonstrated varying connectivity configurations between seven functional domains: subcortical (SC), auditory (AUD), sensorimotor (SMN), visual (VIS), cognitive control (CC), default mode network (DMN), and cerebellar (CB) domains. Darker red color implies stronger positive, darker blue stronger negative connectivity. Stated percentages correspond to state-specific fraction times across all subjects. State 1 was characterized by the most pronounced positive intra- and negative inter-domain connectivity. Subject spent the least amount of time in this state in total. State 2, on the other hand, featured particularly weak intra- and mostly neutral inter-domain connectivity. It was the most frequent state. State 3 delineated positive intra- and importantly also positive inter-domain connectivity between sensorimotor and visual domains. The ordering of states corresponds to the one introduced by the k-means algorithm.

### Temporal characteristics

Next, we evaluated differences in dynamic patterns between the three groups of patients (Stroke severity: mild: 0–2 vs. moderate: 3–9 vs. severe: 10–21 NIHSS). By means of three-level one-way ANOVAs, we detected significant differences of dynamic patterns relating to State 1, the state with the highest functional segregation (Fraction and dwell times of State 1: *p*<0.05, **Figure 3**). While mildly and moderately affected patients did not differ based on post hoc t-tests, we found significantly different fraction times between mildly and severely affected patients as well as significantly different dwell times between mildly and severely as well as moderately and severely affected patients (*p*<0.05, FDR-corrected). Severely affected patients generally spent more time in State 1. These significant differences between discrete groups of mildly, moderately and severely affected patients were also found in continuous Pearson correlations: The NIHSS-score at time of scanning significantly correlated with fraction, as well as dwell times of State 1 (Fraction time State 1 and NIHSS: *r* = 0.49, *p* = 0.001, dwell time & NIHSS: *r* = 0.55, *p*<0.001). This, once again, demonstrated that more profound symptom severity increased the amount of time spent in the densely connected, highly segregated State 1. We did not observe any significant correlations between any of the fraction or dwell times and recovery in stroke severity, or the WMH lesion load (*p*>0.05).

**Figure 3.**
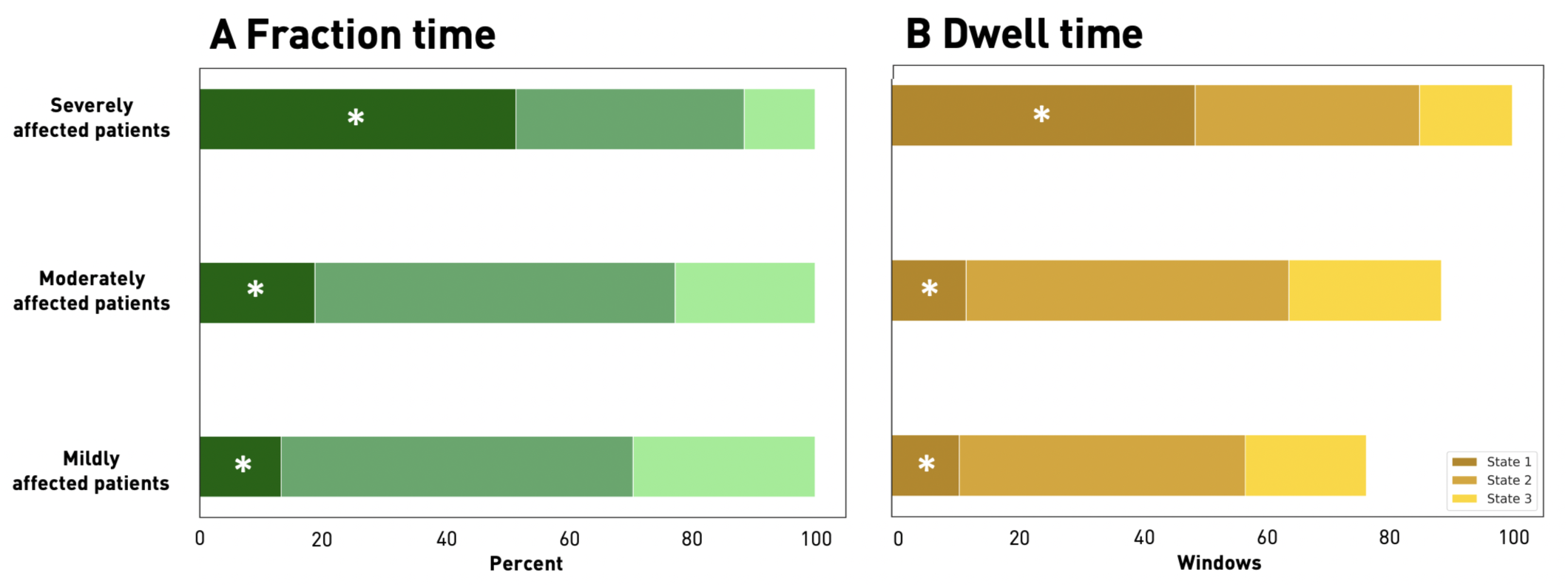
Fraction and dwell times for each of the three dynamic connectivity states and stroke severity defined subgroups of mildly, moderately and severely affected patients (asterisks mark statistically significant differences between patient subgroups based on one-way ANOVAs, *p*<0.05). A Fraction times. Severely affected patients (NIHSS > 9, *upper row*) presented with a markedly different dynamic pattern than moderately (NIHSS 3–9, *middle row*) and mildly (NIHSS < 3, *bottom row*) affected patients: In contrast to the other two patient groups that preferred State 2, a particularly weakly connected state, severely affected patients spent significantly more time in the densely connected State 1. **B Dwell times**. In parallel to the fraction time findings, severely affected patients spent significantly more time in State 1 at any one time in comparison to the less affected patient groups.

### Alterations in dynamic connectivity

Focusing on the differences in dynamic connectivity strengths, we identified numerous connectivity state-specific group differences (one-way ANOVA: *p*<0.05). The densely connected State 1 contained most of these differences (c.f. **Figure 2**). Specifically, post hoc t-tests between mildly and moderately affected patients revealed decreased dynamic connectivity between bilateral precentral areas and the left sensorimotor area (*p*<0.05, FDR-corrected, **Figure 4**, *left panel*). Furthermore, dynamic connectivity was found decreased between bilateral pre- and postcentral sensorimotor areas. Mildly and severely affected patients comprised numerous significantly varying connectivity pairs (post hoc t-tests: *p*<0.05, FDR-corrected, **Figure 4**, *middle panel*). These differences particularly involved connections between the bilateral superior temporal gyri and multiple cortical sensorimotor areas (bilateral ventral precentral areas, left and right sensorimotor areas). Furthermore, a bilateral putamen network comprised several increased connectivity bonds to the already mentioned superior temporal gyrus as well as the left middle temporal gyrus and calcarine gyrus. Lastly, further increased connectivity pairs were found within and between the cognitive control and default mode networks, the visual domain and the bilateral postcentral gyri as well as the cerebellum. Dynamic connectivity differences between moderately and severely affected patients predominantly pertained to bilateral subcortical, superior temporal gyri and ventral precentral networks (post hoc t-tests: *p*<0.05, FDR-corrected, **Figure 4**, *right panel*). State 2, the weakly connected state, encompassed only two dynamic connectivity pairs that were significantly different between patient groups after FDR-correction: Mildly and severely affected patients differed in their dynamic connectivity between the right intraparietal lobule and right middle occipital gyrus. Severely affected patients presented with an increased connectivity between bilateral superior parietal lobules and middle temporal gyri in contrast to moderately affected patients (**Supplementary Figure 2**). State 3 was characterized by increased dynamic connectivity between the bilateral posterior insula and precuneus as well as decreased dynamic connectivity between a bilateral putamen network and anterior insula when contrasting moderately with mildly affected patients. A decreased dynamic connectivity between the lingual gyrus and right middle occipital gyrus was found when comparing severely to mildly affected patients (**Supplementary Figure 3**). Altogether, most of these reported dynamic connectivity pairs were also significantly (Pearson) correlated with NIHSS stroke severity at time of scanning (c.f., **Supplementary Figure 4**). However, such a significant correlation with the NIHSS score at time of scanning was missing for the lateralized dynamic connectivity decrease between cortical motor networks. Consistently, while we observed a significant lateralized connectivity difference between mildly and moderately affected patients in State 1, we did not substantiate such a significant difference, when contrasting mildly or moderately affected with severely affected patients.

**Figure 4.**
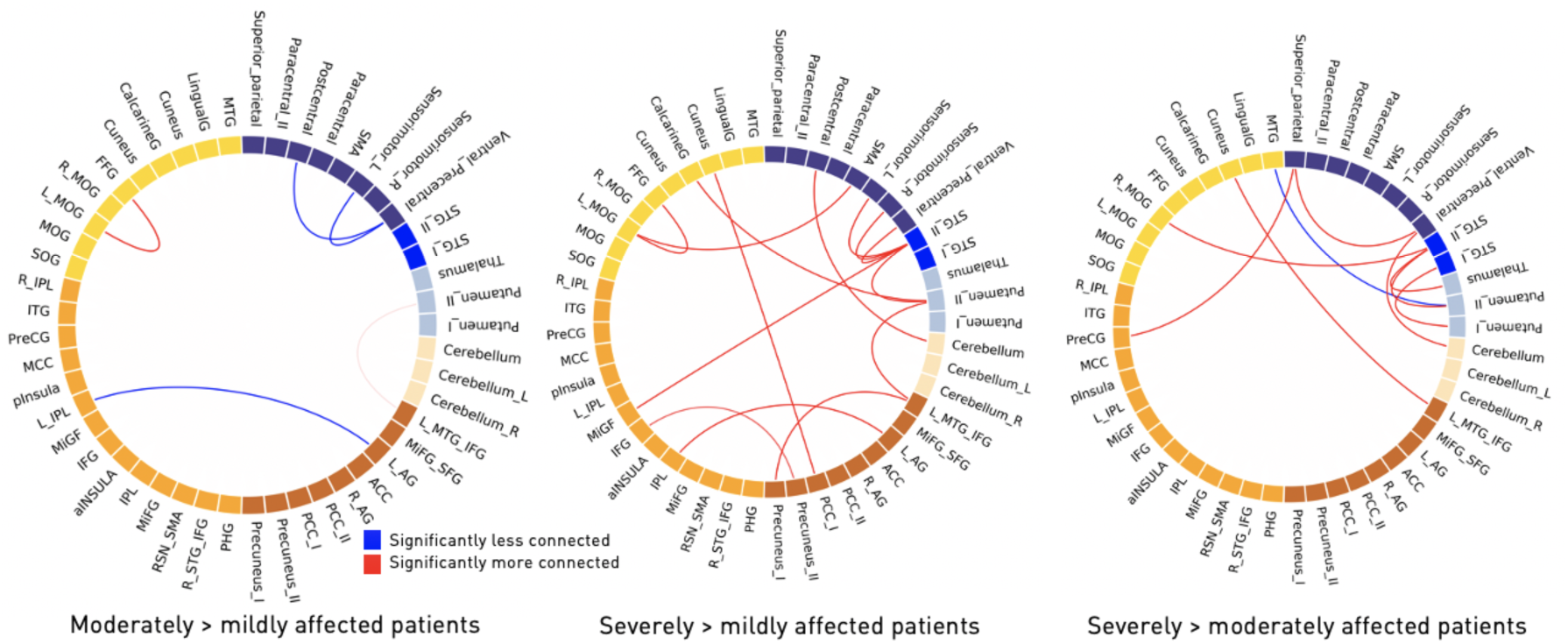
Significant dynamic connectivity differences between mildly, moderately and severely affected patient groups in State 1 (one-way ANOVAs: *p<*0.05, post hoc t-tests: *p<*0.05, FDR-corrected for multiple comparisons). The functionally segregated state 1 comprised the most significantly altered connectivity pairs. Severely affected patients comprised numerous dynamic connectivity pairs with enhanced connectivity compared to both mildly and moderately affected patients. These changes primarily involved subcortical and cortical motor networks, as well as multiple connections to the default mode network.

In a subsequent step, we investigated whether the dynamic connectivity pairs that differed significantly between the three groups also correlated with the realized recovery in stroke severity within the first three months after stroke. We extracted one such connectivity pair with a significant correlation, as well as two connectivity pairs showing strong trends: In State 1, the densely connected state, the connectivity of the bilateral intraparietal lobuley and left angular gyrus was significantly correlated with change in stroke severity (*r* = –0.68, *p*<0.05, FDR-corrected, **Figure 5**, *upper row*). Furthermore, the dynamic connectivity between the bilateral putamen network and superior temporal gyri (STG) as well as the anterior insula within State 3 correlated strongly with the change in stroke severity over time (putamen – STG: *r* = 0.61, *p* = 0.007, uncorrected, *p* = 0.08, FDR-corrected, and putamen – anterior insula: *r* = 0.66, *p* = 0.003, uncorrected, *p* = 0.05, FDR-corrected, **Figure 5B**, *lower row*). The effects of age, sex and motion in the scanner were accounted for in the analyses reported above.

**Figure 5.**
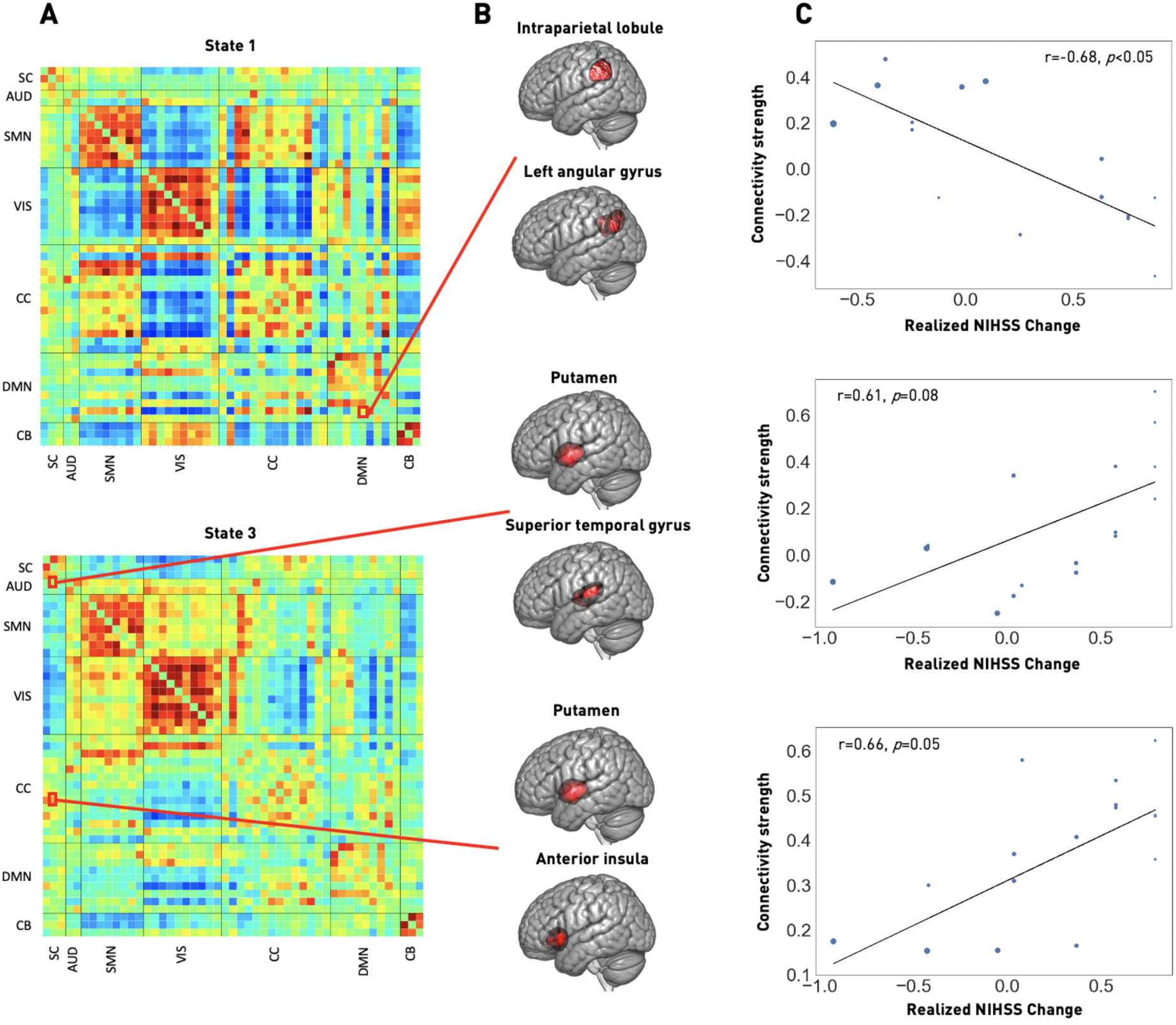
Recovery in the first three months after stroke is linked to acute dynamic connectivity in subcortical (SC), auditory (AUD), cognitive control (CC) and default mode network (DMN) domains. A. Recovery-correlated connectivity pairs are highlighted within dynamic connectivity State 1 (upper row) and State 3 (bottom row). The significantly correlated dynamic connectivity pair in State 1 involved the bilateral intraparietal lobules and left angular gyrus, both located within the DMN domain. In State 3, the dynamic connectivity between the bilateral putamen (SC) and superior temporal gyri (AUD) as well as bilateral putamen and anterior insula (CC) was correlated with the recovery after stroke. **B. Brain renderings of involved networks. C. Correlation plots**. Recovery, measured as realized change in NIHSS and adjusted for the initial NIHSSscan stroke severity, is plotted on the x-axis, dynamic connectivity strength on the y-axis (p-values are FDR-corrected for multiple comparison). The size of the dots corresponds to an individual’s stroke severity at time of scanning.

### Bayesian prediction of acute stroke severity

Bayesian hierarchical models, either taking stroke severity at admission or the three dwell times as input, achieved comparable levels of explained variance when predicting stroke severity at the time of scanning (posterior predictive check: R2-scores: 32.2% and 32.8%, respectively). The joint model combining NIHSSadmission and dwell times increased explained variance to 62.1%. The leave-one-out-cross-validation based Bayesian model comparison also suggested the extended acute model, including both NIHSSadmission and dwell times, as the best performing model (LOOCV-estimated deviance ± standard error (SE): 224.1±11.9). The further two models, considering either NIHSSadmission or the dwell times, followed on par (Deviance ± SE: 242.7±6.6 and 246.2±10.3, respectively). Interpreting parameter weights: A higher NIHSSadmission score predicted a higher NIHSS at time of scanning (posterior mean: 0.51, highest probability density interval (HPDI) covering 94% uncertainty: 0.29–0.74, **Supplementary figure 5A**). The same was true for higher dwell times in State 1 and, to a lesser degree, to higher dwell times in State 2 and 3 (State 1: posterior mean: 0.124, HPDI: 0.063–0.182; State 2: posterior mean: 0.028, HPDI: –0.012 – 0.064; State 3: posterior mean: 0.028, HPDI: –0.023 – 0.080, **Supplementary figure 5B**). Varying intercepts for the groups of moderate and high white matter hyperintensity loads only diverged in case of the model including admission NIHSS only (moderate WMH load: posterior mean: –0.266, HPDI: –3.786 – 3.066, high WMH load: posterior mean: 1.441, HPDI: –0.659 – 3.667). Here, higher white matter hyperintensity load indicated a higher NIHSS outcome at the time of scanning. A similar divergence of intercepts was not visible in the acute dynamic or extended acute models. Ancillary analyses suggested a trend for higher stroke severities with increasing age, but did not suggest any additional effect of the administration of rtPA or sex on the stroke severity at time of scanning. The explained variance increased to 64.9%.

## Discussion

Dynamic functional network connectivity analyses permit the evaluation of brain connectivity alterations in the range of seconds^11,12,13^. This approach, therefore, may capture naturally fluctuating neural signals in a more veridical and behaviorally relevant way^38^. We here examined alterations of whole-brain dynamic connectivity in 41 AIS patients in relation to the severity of their acute stroke deficit. We identified three dynamic connectivity states, which were strongly related to the severity of symptoms. Most remarkably, severely affected stroke patients (NIHSS>9) spent significantly more time in the segregated State 1 with highly positive intra- and highly negative inter-domain connectivity. This strong intra-domain connectivity was particularly apparent for the visual and sensorimotor domains. In contrast, State 2, a weakly connected state, comprised rather low intra- and mostly neutral inter-domain connectivity. State 3 featured a combination of both previous states with positive intra-domain connectivity and slightly positive inter-domain connectivity. While fraction and dwell times for State 2 and 3 did not differ significantly between patient groups, all three states comprised significantly altered dynamic connectivity between multiple brain areas.

## Segregation and stroke severity

Patients experiencing severe strokes had a significant predilection for the densely connected State 1. Since this state exhibited strong positive intra-network connectivity, brain areas belonging to the same functional domain, e.g. the pre- and post-central areas of the cortical sensorimotor domain, could easily exchange information. Conversely, the strong negative inter-network connectivity indicated a lower level of communication between different functional domains. This pattern of isolated information processing within functional domains can be interpreted as *functional segregation. Functional integration*, on the other hand, implies an effortless information transfer within and between functional domains^39,40^.

A comparable preference for a segregated state in case of more severe deficits has recently been described in an independent cohort of acute stroke patients, who presented with a comparable spectrum of symptom severity and were also scanned within the first days after their cerebrovascular events^18^. In this study, patients with severe motor impairments had a significantly higher probability of transitioning into a dynamic connectivity state that was characterized by a high segregation between motor domains. The combination of both studies thus suggests some robustness of these findings.

Further non-stroke-related studies suggest an association between higher levels of segregation and larger gains in cognitive and motor skill learning in health^41,42,43^ and disease^44^ (for a recent review: ^45^). Nonetheless, we here did not find a significant correlation between the level of segregation and changes in stroke severity in the first three months post-stroke. Therefore, the dynamic increase in segregation observed in our study primarily appeared as an expression of deteriorated, lost function, and not a mechanism supporting brain plasticity and recovery.

Neuropsychiatric diseases, such as schizophrenia, bipolar disorder or autism, have primarily been linked to increased fraction and dwell times in weakly connected states, such as our State 2 (c.f., schizophrenia:^25,46^, bipolar disorder:^47^, autism:^48^). This increased occurrence of less segregated states was suggested to represent reduced vigilance and enhanced self-focused thought^12,49^. On the other hand, many neurological diseases, e.g. migraine, Parkinson’s disease and – as also presented here – stroke show the opposite pattern. Neurological patients apparently exhibit a preference for highly connected, segregated dynamic connectivity states, such as our State 1. This preference may thus denote a joint signature of limited neurological function (c.f., migraine:^14^, Parkinson’s disease:^15^, stroke:^18^).

## Alterations in dynamic connectivity: Acute stroke severity

We detected wide-spread differences in dynamic connectivity between brain areas of various functional domains. These connectivity pairs were significantly altered between the three severity defined groups of stroke patients and also significantly correlated with stroke severity at time of scanning across the entirety of stroke patients. We observed the previously described inter-hemispheric decrease in connectivity between cortical motor areas^5,6,7,8^ only between mildly and moderately affected patients. However, similar alterations were not detectable between mildly and severely or moderately and severely affected patients. A recent study reported comparable findings: When contrasted with healthy controls, stroke patients with moderate upper limb impairments expressed more pronounced decreases in interhemispheric dynamic connectivity than did severely affected patients^18^. Further previous studies had defined this specific decrease between bilateral motor areas based on comparisons between stroke patients and controls^5,6,7,8^. However, these studies had not supplemented any more granular analyses between connectivity strength and the amount of impairment. Thus, future studies could aim to confirm and further elucidate the biological meaning of bilateral motor area connectivity in relation to stroke severity – e.g. is it beneficial or detrimental in the process of stroke recovery?

## Alterations in dynamic connectivity: Recovery of stroke severity

Dynamic connectivity between three network pairs was either significantly correlated with the realized recovery in stroke severity in the first three months after stroke or showed strong trends. The densely connected State 1 comprised a significant negative correlation between the change in stroke severity and the dynamic connectivity of the default mode networks *bilateral intraparietal lobule* and *left angular gyrus*. Additionally, State 3 presented two positive correlations between recovery in stroke severity and the dynamic connectivity of *bilateral putamen* and *bilateral superior temporal*, as well as the same *putamen network* and *bilateral anterior insula*. Observed changes hence involved the subcortical motor, auditory, cognitive control and default mode network domains. The revealed importance of the subcortical putamen connectivity for stroke outcome may primarily relate to the recovery of motor symptoms. With regard to the cognitive control and default mode networks: These domains are well known to encompass cortical hubs that are globally well-connected and essential to orchestrate various brain functions^50,51,52^. Recent stroke studies relying on structural MRI data already indicated worse stroke outcomes the more of such well-connected areas were affected by stroke lesions^36,53^. Previous functional MRI studies have also reported disturbed functional connectivity within the default mode network when comparing subacute ischemic stroke patients and healthy controls, independent of any specific deficits or stroke severity (^54^: decreased left medial temporal lobe, posterior cingulate and medial prefrontal cortical areas,^10^: decreased posterior cingulate cortex/precuneus, increased medial prefrontal cortex, left hippocampus). While the extracted areas slightly differ between studies, likely due to varying outcome measures, group definitions and scanning time points, previous studies and ours combined suggest that cognitive control and default mode networks may play essential roles for stroke outcome and recovery in addition to motor-related domains.

## Modelling stroke severity

The individual dwell times in all of the three states proved to be equally effective in predicting the stroke severity at the time of scanning as the NIHSS score at the time of admission based on our Bayesian hierarchical models^35^. Moreover, the combination of dynamic connectivity estimates and clinical information led to the highest prediction performance and also the highest-ranked model in the Bayesian model comparison. However, the maximum explained variance of 62% still leaves a substantial amount of the variance in recovery unexplained and might thus call for an even more comprehensive collection of outcome predictors^55^.

Nonetheless, the increase in prediction performance based on our dynamic connectivity estimates can be seen as evidence, that these estimates represent valid predictors of stroke outcome. This is yet a further demonstration of the potential utility of neuroimaging markers for the prediction of acute stroke symptoms. Previous studies have already begun to highlight the value of MRI derived estimates. Examples of such predictors comprise the number of lesioned rich club regions, as obtained from structural MRI data^53^, lesion topographies as inferred from acute diffusion tensor imaging^56^, static connectivity between motor areas^8^ and information on the network topology^36^, as characterized by functional MRI data.

White matter disease has been associated with poorer early neurological outcomes after stroke in recent years^57,58^. We here investigated the differential effects of white matter hyperintensities on stroke severity by means of a hierarchical intercept term. In case of the admission-NIHSS model, group-wise intercepts diverged: In accordance with previous studies, a higher white matter hyperintensity lesion load was predictive of a higher stroke severity at the time of scanning. Importantly, this effect was independent of the initial admission stroke severity.

## Further limitations and future directions

While of relatively modest sample size (n = 41), this study is both unique to and yet comparable with other published stroke cohorts undergoing rsfMRI in the acute post-stroke recovery phase. It is unique in its approach of assessing acute alterations in dynamic connectivity exclusively within strata of variably affected stroke patients, without the direct comparison to a healthy control group. Furthermore, it is comparable in size with prior dynamic functional connectivity studies in the first few days after stroke (^59^: 19 patients;^18^: 31 patients). Notably, one of our main findings, i.e. favored transiently increased segregation in case of a high stroke severity, is well in line with previous reports of increased transition likelihood to segregated states in case of severe motor impairments^18^. Altogether, this agreement demonstrates the overall robustness of results for these kinds of dataset sizes.

In this study, we focused on global stroke severity, measured on the NIHSS scale, as the main outcome of interest. The NIHSS can already be considered more granular than the frequently used modified Rankin Scale score. Nonetheless, the unavailability of an even more fine-grained score that would capture specific impairments, e.g., in the cognitive or language domains, can be seen as a current limitation. Especially in view of numerous insight-generating dynamic functional network connectivity studies in the field of cognitive decline (e.g., subcortical ischemic vascular disease:^60^Alzheimer’s disease:^61,62^), future studies are warranted to explore dynamic connectivity alterations in relation to these specific symptoms post-stroke further. Lastly, we only recruited stroke patients with higher WMH lesions loads. This criterion may have had an effect on functional connectivity on its own, as previous literature suggests connectivity alterations based on white matter disease^63,64^. However, we here did not find any significant correlations between the WMH lesion load and any of the dynamic connectivity estimates. We furthermore included the WMH lesion load in our Bayesian prediction models, which rendered their influence on stroke outcome apparent and may motivate a more frequent inclusion as proxy of chronic small vessel disease in future stroke imaging studies.

## Conclusion

We here revealed transiently increased segregation in severe stroke by leveraging dynamic functional network connectivity analyses. Since we did not observe a correlation between segregation and the amount of recovery post-stroke, the enhanced moment-to-moment segregation was primarily interpretable as expression of deteriorated function. The change in stroke severity in the first three months post-stroke was furthermore linked to dynamic connectivity involving default mode network components, suggesting a pivotal role of this domains in stroke recovery.

## Data Availability

Scripts used for the generation of dynamic connectivity results will be made available upon publication.

## Acknowledgements

We are grateful to our colleagues at the J. Philip Kistler Stroke Research Center for valuable support and discussions. Furthermore, we are grateful to our research participants without whom this work would not have been possible.

## Supplementary Material

**Supplementary figure 1.**
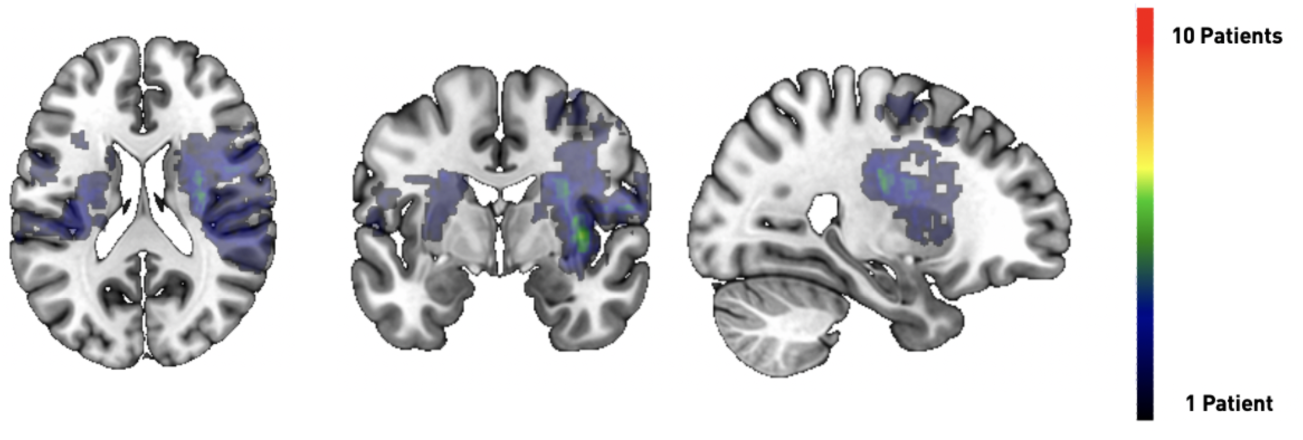
Lesion overlap.

**Supplementary figure 2.**
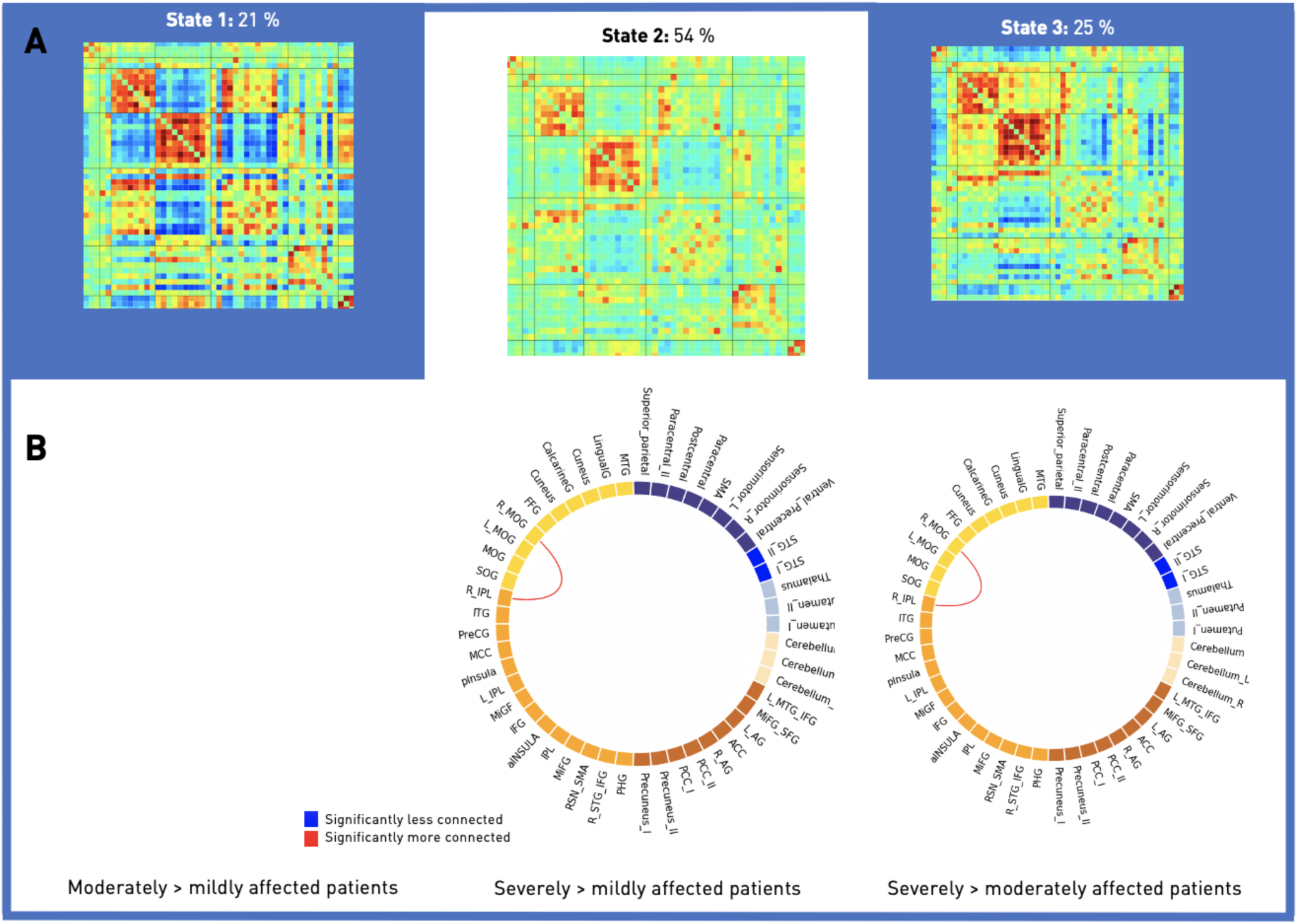
Significantly altered dynamic connectivity pairs in State 2 (post-hoc t-tests: *p*<0.05, FDR-corrected for multiple comparison).

**Supplementary figure 3.**
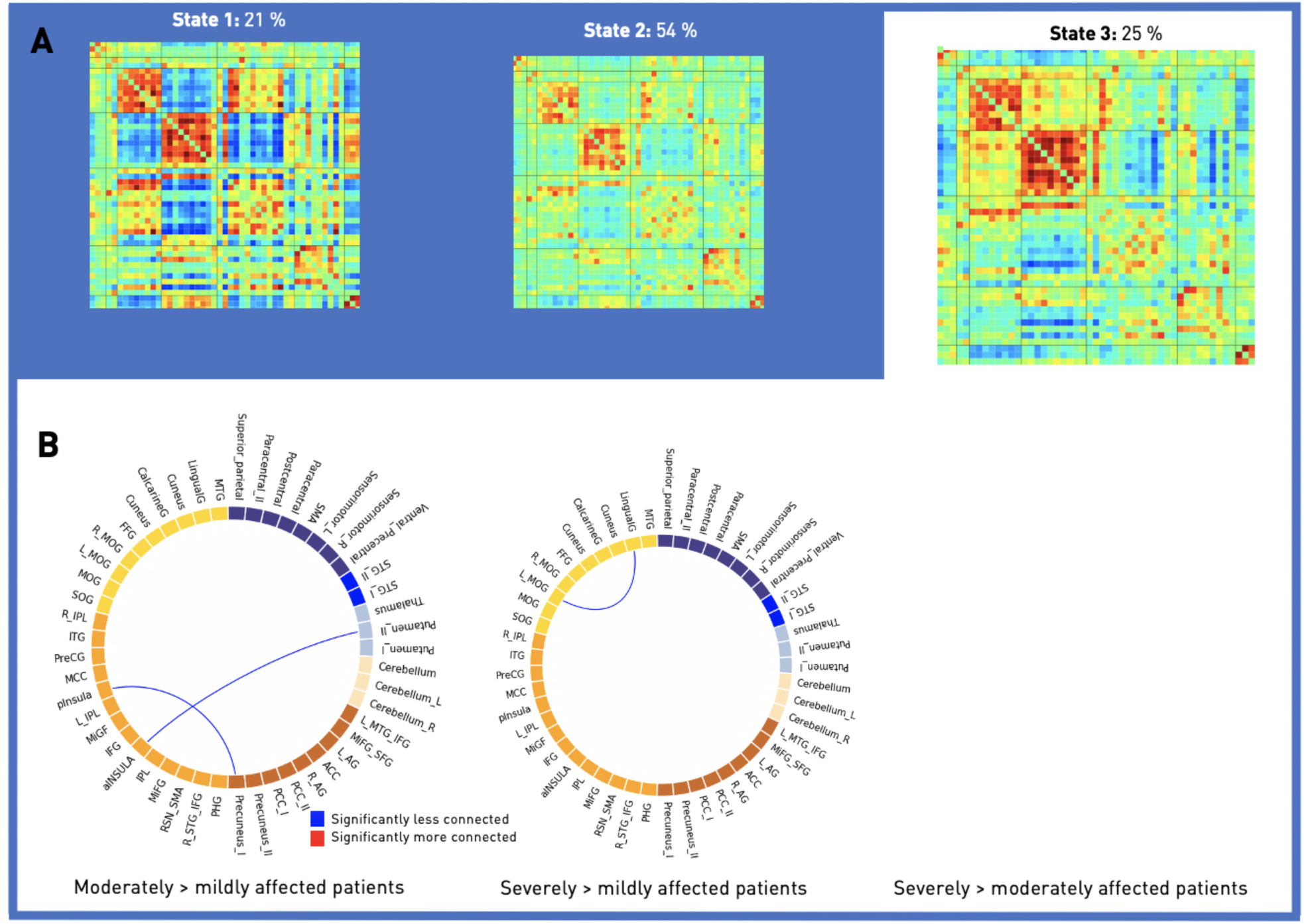
Significantly altered dynamic connectivity pairs in State 3 (post-hoc t-tests: *p*<0.05, FDR-corrected for multiple comparison).

**Supplementary figure 4.**
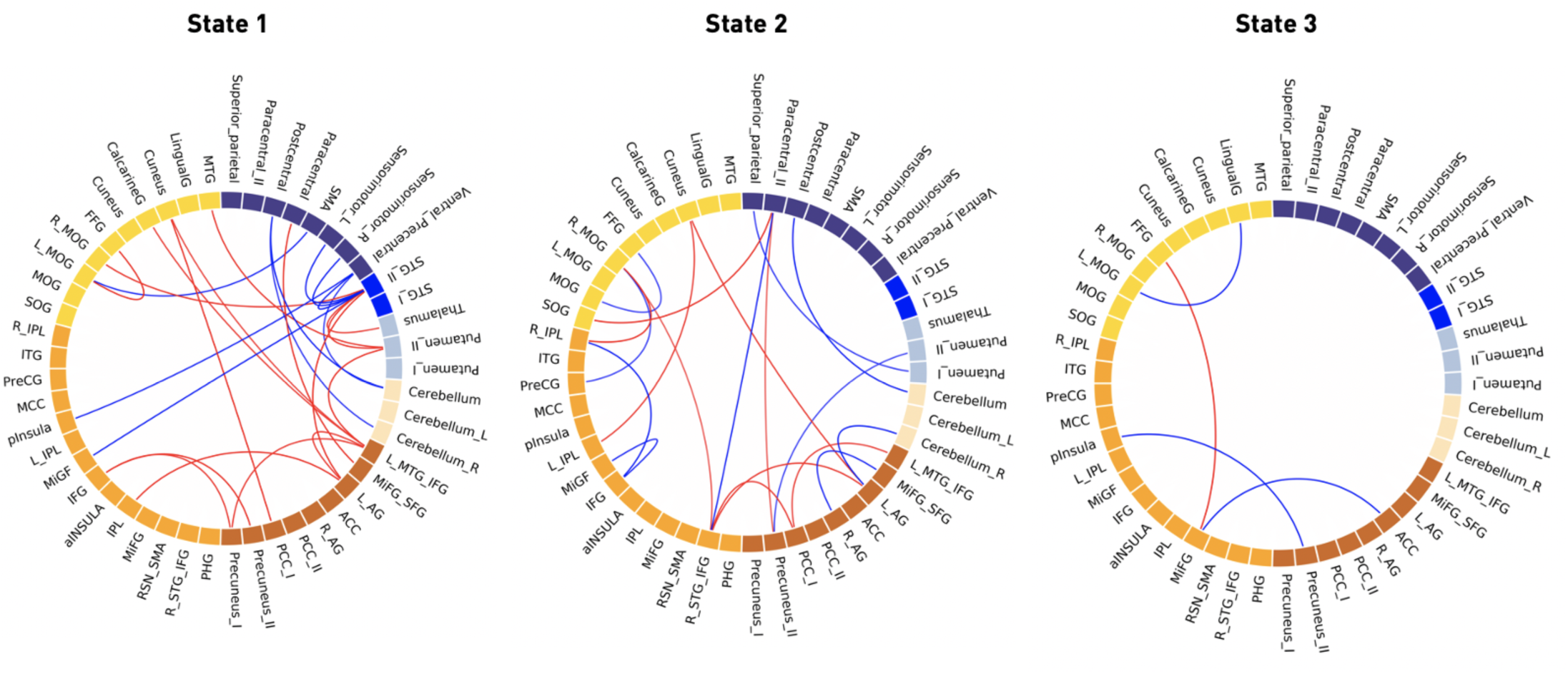
Dynamic connectivity pairs significantly correlated with stroke severity at the time of scanning (*p*<0.05, FDR-corrected for multiple comparison).

**Supplementary figure 5.**
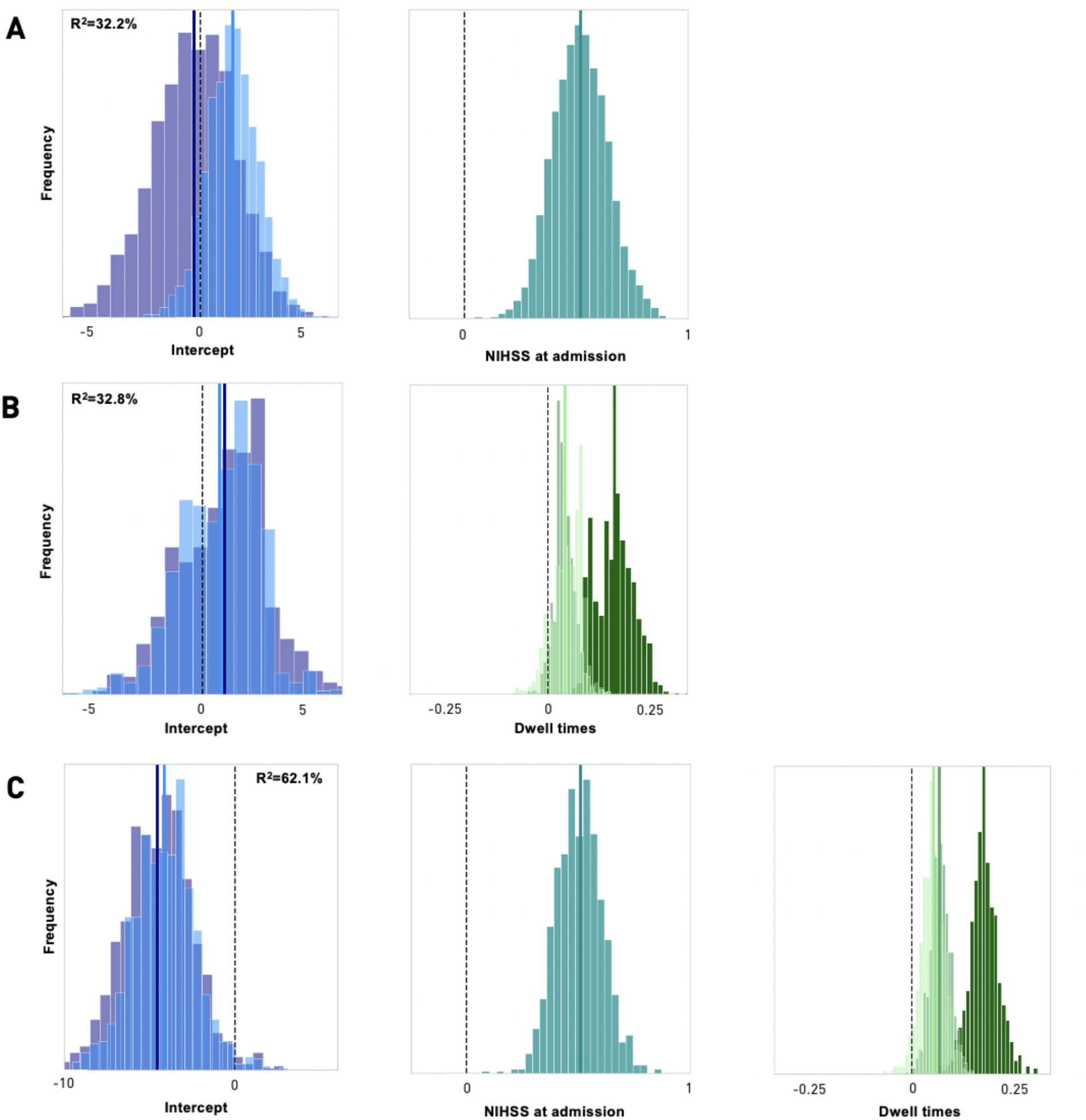
Bayesian hierarchical modelling of the stroke severity at time of scanning: Posterior parameter distributions. **A. Acute baseline model.** The model based on the NIHSS score at admission, thus on average 3 days earlier, could predict the NIHSS at time of scanning with an explained variance of 32.3% (obtained via posterior predictive checks). The intercept for patients with a higher white matter hyperintensity load indicated a higher predicted NIHSS score at time of scanning (*lightblue*) compared to the group of patients with a lower white matter hyperintensity load (*darkblue*). The parameter posterior mean of 0.51 for the NIHSS scores at admission denoted a decrease in NIHSS stroke severity until the time of scanning (right plot). **B. Acute dynamic model**. A higher dwell time in any of the three states predicted a higher NIHSS score at the time of scanning, the explained variance was 32.8%. This effect was particularly strong for dwell times in State 1 (*darkgreen*). The effect of the white matter hyperintensity load on stroke severity did not differ between the groups of moderate and high white matter hyperintensity loads. **C. Acute extended model relying on the NIHSS at admission as well as the derived dwell times**. A higher NIHSS score at admission, as well as higher dwell times, mainly in State 1, continued to be predictive of a higher NIHSS score at the time of scanning. Explained variance of the joint model was 62.1%.

## References

1. Collaborators, G. 2016 L. R. of S. Global, Regional, and Country-Specific Lifetime Risks of Stroke, 1990 and 2016. New England Journal of Medicine 379, 2429–2437 (2018).

2. Corbetta, M. et al. Common Behavioral Clusters and Subcortical Anatomy in Stroke. Neuron 85, 927–941 (2015).

3. Grefkes, C. & Fink, G. R. Connectivity-based approaches in stroke and recovery of function. The Lancet Neurology 13, 206–216 (2014).

4. Ward, N. S. Restoring brain function after stroke—bridging the gap between animals and humans. Nature Reviews Neurology 13, 244 (2017).

5. Carter, A. R. et al. Resting interhemispheric functional magnetic resonance imaging connectivity predicts performance after stroke. Annals of neurology 67, 365–375 (2010).

6. Wang, L. et al. Dynamic functional reorganization of the motor execution network after stroke. Brain 133, 1224–1238 (2010).

7. Golestani, A.-M., Tymchuk, S., Demchuk, A., Goodyear, B. G. & VISION-2 Study Group. Longitudinal Evaluation of Resting-State fMRI After Acute Stroke With Hemiparesis. Neurorehabilitation and Neural Repair 27, 153–163 (2013).

8. Rehme, A. K. et al. Identifying neuroimaging markers of motor disability in acute stroke by machine learning techniques. Cerebral cortex 25, 3046–3056 (2014).

9. Lassalle-Lagadec, S. et al. Subacute Default Mode Network Dysfunction in the Prediction of Post-Stroke Depression Severity. Radiology 264, 218–224 (2012).

10. Ding, X. et al. Patterns in default-mode network connectivity for determining outcomes in cognitive function in acute stroke patients. Neuroscience 277, 637–646 (2014).

11. Chang, C. & Glover, G. H. Time–frequency dynamics of resting-state brain connectivity measured with fMRI. Neuroimage 50, 81–98 (2010).

12. Allen, E. A. et al. Tracking Whole-Brain Connectivity Dynamics in the Resting State. Cerebral Cortex 24, 663–676 (2014).

13. Calhoun, V. D., Miller, R., Pearlson, G. & Adalı, T. The Chronnectome: Time-Varying Connectivity Networks as the Next Frontier in fMRI Data Discovery. Neuron 84, 262–274 (2014).

14. Tu, Y. et al. Abnormal thalamocortical network dynamics in migraine. Neurology 92, e2706–e2716 (2019).

15. Kim, J. et al. Abnormal intrinsic brain functional network dynamics in Parkinson’s disease. Brain 140, 2955–2967 (2017).

16. Fiorenzato, E. et al. Dynamic functional connectivity changes associated with dementia in Parkinson’s disease. Brain (2019).

17. Espinoza, F. A. et al. Whole-Brain Connectivity in a Large Study of Huntington’s Disease Gene Mutation Carriers and Healthy Controls. Brain Connectivity 8, 166–178 (2018).

18. Bonkhoff, A. K. et al. Acute ischaemic stroke alters the brain’s preference for distinct dynamic connectivity states. Brain (2020) doi:10.1093/brain/awaa101.

19. Fazekas, F., Chawluk, J. B., Alavi, A., Hurtig, H. I. & Zimmerman, R. A. MR signal abnormalities at 1.5 T in Alzheimer’s dementia and normal aging. American journal of roentgenology 149, 351–356 (1987).

20. Ashburner, J. & Friston, K. J. Unified segmentation. Neuroimage 26, 839–851 (2005).

21. Du, Y. & Fan, Y. Group information guided ICA for fMRI data analysis. Neuroimage 69, 157–197 (2013).

22. Salman, M. S. et al. Group ICA for identifying biomarkers in schizophrenia:‘Adaptive’networks via spatially constrained ICA show more sensitivity to group differences than spatio-temporal regression. NeuroImage: Clinical 22, 101747 (2019).

23. Cox, R. W. AFNI: software for analysis and visualization of functional magnetic resonance neuroimages. Computers and Biomedical research 29, 162–173 (1996).

24. Rachakonda, S., Egolf, E., Correa, N. & Calhoun, V. Group ICA of fMRI toolbox (GIFT) manual. *Dostupné z http://www.nitrc.org/docman/view.php/55/295/v1.3d_GIFTManualpdf* [*cit. 2011-11-5*] (2007).

25. Damaraju, E. et al. Dynamic functional connectivity analysis reveals transient states of dysconnectivity in schizophrenia. NeuroImage: Clinical 5, 298–308 (2014).

26. Sakoğlu, Ü. et al. A method for evaluating dynamic functional network connectivity and task-modulation: application to schizophrenia. Magnetic Resonance Materials in Physics, Biology and Medicine 23, 351–366 (2010).

27. Friedman, J., Hastie, T. & Tibshirani, R. Sparse inverse covariance estimation with the graphical lasso. Biostatistics 9, 432–441 (2008).

28. Varoquaux, G., Gramfort, A., Poline, J.-B. & Thirion, B. Brain covariance selection: better individual functional connectivity models using population prior. in Advances in neural information processing systems 2334–2342 (2010).

29. Smith, S. M. et al. Network modelling methods for FMRI. NeuroImage 54, 875–891 (2011).

30. Lloyd, S. Least squares quantization in PCM. IEEE transactions on information theory 28, 129–137 (1982).

31. Aggarwal, C. C., Hinneburg, A. & Keim, D. A. On the surprising behavior of distance metrics in high dimensional space. in International conference on database theory 420–434 (Springer, 2001).

32. Hutchison, R. M. et al. Dynamic functional connectivity: Promise, issues, and interpretations. NeuroImage 80, 360–378 (2013).

33. Espinoza, F. A. et al. Dynamic functional network connectivity in Huntington’s disease and its associations with motor and cognitive measures. Human Brain Mapping (2019) doi:10.1002/hbm.24504.

34. Lin, D. J. et al. Corticospinal tract injury estimated from acute stroke imaging predicts upper extremity motor recovery after stroke. Stroke 50, 3569–3577 (2019).

35. Gelman, A. & Hill, J. Data analysis using regression and multilevel/hierarchical models. (Cambridge university press, 2006).

36. Ktena, S. I. et al. Brain connectivity measures improve modeling of functional outcome after acute ischemic stroke. Stroke 50, 2761–2767 (2019).

37. Salvatier, J., Wiecki, T. V. & Fonnesbeck, C. Probabilistic programming in Python using PyMC3. PeerJ Computer Science 2, e55 (2016).

38. Vidaurre, D., Arenas, A. L., Smith, S. M. & Woolrich, M. W. Behavioural relevance of spontaneous, transient brain network interactions in fMRI. bioRxiv 779736 (2019).

39. Friston, K. J. Functional and Effective Connectivity: A Review. Brain Connectivity 1, 13–36 (2011).

40. Eickhoff, S. B. & Grefkes, C. Approaches for the integrated analysis of structure, function and connectivity of the human brain. Clinical EEG and neuroscience 42, 107–121 (2011).

41. Gallen, C. L. et al. Modular brain network organization predicts response to cognitive training in older adults. PloS one 11, e0169015 (2016).

42. Baniqued, P. L. et al. Brain network modularity predicts exercise-related executive function gains in older adults. Frontiers in aging neuroscience 9, 426 (2018).

43. Mattar, M. G. et al. Predicting future learning from baseline network architecture. Neuroimage 172, 107–117 (2018).

44. Arnemann, K. L. et al. Functional brain network modularity predicts response to cognitive training after brain injury. Neurology 84, 1568–1574 (2015).

45. Gallen, C. L. & D’Esposito, M. Brain modularity: A biomarker of intervention-related plasticity. Trends in cognitive sciences (2019).

46. Du, Y. et al. Artifact removal in the context of group ICA: A comparison of single‐subject and group approaches. Human brain mapping 37, 1005–1025 (2016).

47. Rashid, B., Damaraju, E., Pearlson, G. D. & Calhoun, V. D. Dynamic connectivity states estimated from resting fMRI Identify differences among Schizophrenia, bipolar disorder, and healthy control subjects. Frontiers in human neuroscience 8, 897 (2014).

48. Fu, Z. et al. Transient increased thalamic-sensory connectivity and decreased whole-brain dynamism in autism. NeuroImage 190, 191–204 (2019).

49. Marusak, H. A. et al. Dynamic functional connectivity of neurocognitive networks in children. Human brain mapping 38, 97–108 (2017).

50. Hagmann, P. et al. Mapping the Structural Core of Human Cerebral Cortex. PLoS Biology 6, e159 (2008).

51. Cole, M. W., Pathak, S. & Schneider, W. Identifying the brain’s most globally connected regions. NeuroImage 49, 3132–3148 (2010).

52. Van Den Heuvel, M. P. & Sporns, O. Rich-club organization of the human connectome. Journal of Neuroscience 31, 15775–15786 (2011).

53. Schirmer, M. D. et al. Rich-Club Organization: An Important Determinant of Functional Outcome After Acute Ischemic Stroke. Front. Neurol. 10, (2019).

54. Tuladhar, A. M. et al. Default Mode Network Connectivity in Stroke Patients. PLoS One 8, (2013).

55. Bonkhoff, A. K. et al. Bringing Proportional Recovery into Proportion: Bayesian Hierarchical Modelling of Post-Stroke Motor Performance. medRxiv 19009159 (2019).

56. Moulton, E., Valabregue, R., Lehéricy, S., Samson, Y. & Rosso, C. Multivariate prediction of functional outcome using lesion topography characterized by acute diffusion tensor imaging. NeuroImage: Clinical 23, 101821 (2019).

57. Etherton, M. R., Wu, O. & Rost, N. S. Recent Advances in Leukoaraiosis: White Matter Structural Integrity and Functional Outcomes after Acute Ischemic Stroke. Current Cardiology Reports 18, (2016).

58. Etherton, M. R. et al. White matter integrity and early outcomes after acute ischemic stroke. Translational stroke research 10, 630–638 (2019).

59. Hu, J. et al. Dynamic Network Analysis Reveals Altered Temporal Variability in Brain Regions after Stroke: A Longitudinal Resting-State fMRI Study. Neural Plasticity 2018, 1–10 (2018).

60. Fu, Z. et al. In search of multimodal brain alterations in Alzheimer’s and Binswanger’s disease. NeuroImage: Clinical 101937 (2019) doi:10.1016/j.nicl.2019.101937.

61. Jones, D. T. et al. Non-Stationarity in the “Resting Brain’s” Modular Architecture. PLoS ONE 7, e39731 (2012).

62. de Vos, F. et al. A comprehensive analysis of resting state fMRI measures to classify individual patients with Alzheimer’s disease. NeuroImage 167, 62–72 (2018).

63. Schaefer, A. et al. Early small vessel disease affects frontoparietal and cerebellar hubs in close correlation with clinical symptoms—a resting-state fMRI study. Journal of Cerebral Blood Flow & Metabolism 34, 1091–1095 (2014).

64. Reijmer, Y. D. et al. Decoupling of structural and functional brain connectivity in older adults with white matter hyperintensities. Neuroimage 117, 222–229 (2015).

